# Effectiveness of simplifying antiretroviral therapy to maintain viral suppression and improve bone and renal health: comparing simplified and non-simplified therapy

**DOI:** 10.1101/2025.03.20.25324347

**Authors:** Juliana Olsen Rodrigues, Alexandre Naime Barbosa, Stephanie Valentini Ferreira Proença, Lenice Rosário de Souza

**Affiliations:** UNESP (Sao Paulo State University), Infectology Department, Botucatu, SP, Brazil

**Keywords:** HIV, tenofovir, osteopenia, simplification, glomerular filtration rate

## Abstract

**Objective:** Nucleoside/nucleotide reverse transcriptase inhibitors, particularly tenofovir, can cause long-term side effects such as decreased bone mineral density and estimated glomerular filtration rate. A strategy to mitigate these effects is the simplification of antiretroviral therapy, which involves withdrawing one of the nucleoside/nucleotide reverse transcriptase inhibitors from the therapeutic scheme. While clinical trials and real-world studies have demonstrated that the simplified therapy maintains undetectable viral loads, its impact on bone mineral density and kidney function remains unclear owing to the lack of real-world evidence.

**Methods:** This retrospective cohort study compared 152 patients who underwent antiretroviral therapy simplification (primarily due to osteopenia, osteoporosis, or decreased estimated glomerular filtration rate) with 306 patients who maintained triple therapy, between April 2013 and September 2022. The simplified regimens included lamivudine plus dolutegravir or ritonavir-boosted darunavir. The groups were analyzed based on their demographic characteristics using *Student’s t-test* in the case of symmetric data. Therapeutic success (undetectable viral load at the end of follow-up) was assessed using *Kaplan Meier survival analysis*. The estimated glomerular filtration rate variation before and after simplification was analyzed using the *Mann-Whitney test*. Pre-and post-simplification bone mineral density values were evaluated using the chi-square test for trends and assessed in the simplified therapy group. A significance level of 5% (α = 0.05) was adopted for all tests.

**Results:** Simplified antiretroviral therapy was non-inferior to triple therapy in maintaining undetectable viral load. Patients receiving simplified regimens showed a positive variation in estimated glomerular filtration rate. A small subset of patients also exhibited improvements in bone mineral density after antiretroviral therapy simplification.

**Conclusions:** These findings suggest that simplified therapy is as effective as triple therapy and has the additional benefit of reducing tenofovir-related adverse events.

## 1. Introduction

In June 1981, the first cases of acquired immunodeficiency syndrome (AIDS) were described by the *Centers for Disease Control and Prevention*, in men who have sex with men, in New York and Los Angeles, USA. This was the beginning of the human immunodeficiency virus (HIV)/AIDS pandemic, which has resulted in more than 84 million infections and 40 million deaths worldwide in the last 40 years [1,2]. At the beginning of the pandemic, illness and death from HIV/AIDS were almost inevitable within approximately 8 to 10 years after infection because the drugs available at the time were incapable of achieving sustained virological suppression [2,3]. Highly active antiretroviral therapy (HAART), introduced in 1996, containing three active drugs from at least two different classes, significantly reduced the risk of developing AIDS and the number of deaths from the disease. In the last two decades, HAART has been able to maintain viral suppression in the long term, and as a result, the life expectancy of people living with HIV (PLHIV) is currently very similar to that of the general population [1–3].

Healthy aging has become a major goal in the treatment of PLHIV. Complications and comorbidities not associated with the virus, such as cardiovascular, bone, and kidney diseases, are currently more important causes of morbidity in PLHIV than opportunistic infections [4]. Tenofovir (TDF) is one of the drugs of choice for starting HAART and is associated with nephrotoxicity and decreased bone mineral density (BMD) [5]. Nephrotoxicity is related to an accelerated decline in glomerular filtration rate (GFR) and proximal tubular dysfunction [6–8]. Estimated glomerular filtration rate (eGFR) should be measured every six months in PLHIV who are stable using the *Chronic Kidney disease Epidemiology Collaboration* (CKD-EPI) formula, which is based on serum creatinine levels. Current guidelines for the treatment of PLHIV recommend screening for osteoporosis or osteopenia in postmenopausal women and men over 40 years of age [9]. Some studies have suggested that PLHIV may be at risk of osteopenia/osteoporosis if they have been exposed to TDF for more than five years [10–11].

Given the need to maintain treatment for life to prevent disease progression and reduce the risk of morbidity and mortality, simplified antiretroviral therapy (ART) options are being studied to minimize toxicity and maintain treatment effectiveness. Current HIV treatment guidelines recommend regimens consisting of two nucleoside/nucleotide analog reverse transcriptase inhibitors (NRTIs), as the *backbone*, combined with a third agent that may comprise a non-nucleoside analog reverse transcriptase inhibitor (NNRTI), a protease inhibitor boosted with ritonavir (PI/r), or an integrase inhibitor [1–3].

Strategies for simplification include therapies with two classes of antiretrovirals that have been proven effective in over 95% of patients in several randomized clinical trials (RCTs) demonstrating the efficacy of dual therapy: PI/r + lamivudine (3TC) [7] and 3TC + dolutegravir (DTG) [8].

Real-life observational studies remain scarce; however, they are emerging in the same direction, proving the effectiveness of simplified schemes based on PI/r + 3TC [9,10] and DTG + 3TC [11,12]. In these studies, virological failure rates were less than 10.0% (similar to that in RCTs) in analysis periods ranging from two to three years.

Before simplification, a review of the patient’s ART history and previous virological failures should be carried out by analyzing the presence of resistance mutations in genotyping tests, drug interactions, and the patient’s desire to change therapy, because adherence must be excellent in these cases. Furthermore, the main antiretroviral agent in the simplified therapy must have high potency and genetic barrier [11].

In Brazil, 3TC combined with DTG or darunavir boosted with ritonavir (DRV/r) were evaluated for ART simplification strategies, as they have a high genetic barrier and demonstrate safety and efficacy in maintaining virological suppression [12].

Most patients who are candidates for simplification already have an injury related to the long-term use of TDF, such as osteopenia/osteoporosis or decreased renal function. Assuming that withdrawing TDF from the regimen would improve these lesions, simplification would be an optimal treatment strategy because other NRTI options for maintaining triple therapy may have serious side effects.

The objectives of this study were to demonstrate the non-inferiority of simplified ART with 3TC + DTG and 3TC + DRV/r in maintaining undetectable VL compared to triple therapy, and to determine whether simplification of the scheme improves glomerular filtration rate and BMD.

## 2. Materials and Methods

This study was conducted at the Specialized Ambulatory Service of Infectology, located in a city in the interior of São Paulo, Brazil, using a paired, mixed, concurrent, and nonconcurrent cohort design.

Two groups were retrospectively compared: 153 PLHIV who were on regular ART and transitioned to a simplified treatment regimen (3TC combined with either DTG or DRV/r), and 306 PLHIV selected in a 2:1 ratio during the same period and at the same center. The latter group continued the triple ART regimen consisting of one of the following combinations: 2 NRTIs + 1 NNRTI, 2 NRTIs + 1 PI/r, or 2 NRTIs + 1 INI. Patients aged 18 years or older with undetectable VL were included in this study.

Clinical, antiretroviral, demographic, and virological data from both groups were collected between April 2013 and September 2022 from electronic medical records. These records were accessed from March 2021 to October 2022 using patients’ medical record numbers.

One patient whose regimen was simplified to DTG + DRV/r was excluded from the analysis. Consequently, the total number of participants in the simplified group was adjusted to 152.

As a non-interventional study, the decision to simplify the ART regimen for each participant was determined by attending physicians. The main criteria for simplifying ART were the presence of osteopenia or osteoporosis on dual-energy X-ray absorptiometry (DXA) and/or an eGFR < 75 mL/min/1.73 m^2^. Simplification was also performed in patients with side effects related to other NRTIs, such as anemia and lipodystrophy associated with AZT, increased cardiovascular risk with abacavir (ABC) use, or discontinuation of didanosine (ddI), an NRTI that was withdrawn from the therapeutic arsenal for HIV in Brazil in 2016. The exclusion criteria were previous virological failure, presence of mutations in previous genotyping tests, and chronic hepatitis B virus infection.

The eGFR was calculated based on serum creatinine levels using the CKD-EPI equation [13] in both groups at the beginning and end of follow-up. However, BMD was evaluated pre- and post-simplification in patients who received combined therapy with TDF for more than five years, men over 40 years of age and postmenopausal women in group 1 only.

The lumbar spine and femoral neck were evaluated using the DXA system. This examination is routinely performed at our facility for patients who have been receiving combined therapy with TDF for more than five years, as well as for men over 40 years of age and postmenopausal women. BMD results were classified based on T-scores, as follows: greater than –1.5 indicates normal BMD; between –1.5 and –2.5 indicates osteopenia; and less than –2.5 indicates osteoporosis. The results were interpreted according to the “Consensus of the Brazilian Society of Clinical Densitometry of 2008” [14].

### 2.1 Statistical analysis

Frequencies and percentages were computed for categorical variables. These analyses were stratified into two groups: simplified and non-simplified. Therapeutic success (undetectable VL at the end of follow-up) was assessed using *Kaplan Meier survival analysis (log rank* statistics).

Comparisons of means between groups for quantitative variables were performed using *Student’s t-test* in the case of symmetric data. In the case of asymmetry, gamma distribution adjustments were used for comparisons. The eGFR variation before and after simplification was analyzed using the *Mann-Whitney test*. Pre-and post-simplification BMD values were associated with the chi-square test for trend. A significant level of 5% (α = 0.05) was adopted for all tests, or the corresponding p-value was considered. All analyses were conducted using SAS for Windows (version 9.4) and SPSS 27 (IBM, Armonk, NY, USA).

## 3. Results

No differences were observed between groups; among the 306 patients in the non-simplified group, 298 (97.4%) remained with undetectable VL at the end of follow-up, and the same occurred in 148 (95.4%) patients in the simplified group (p = 0.499). According to Figure 1, the permanence of patients with undetectable VL over time was similar between the two groups.

**Figure 1.**
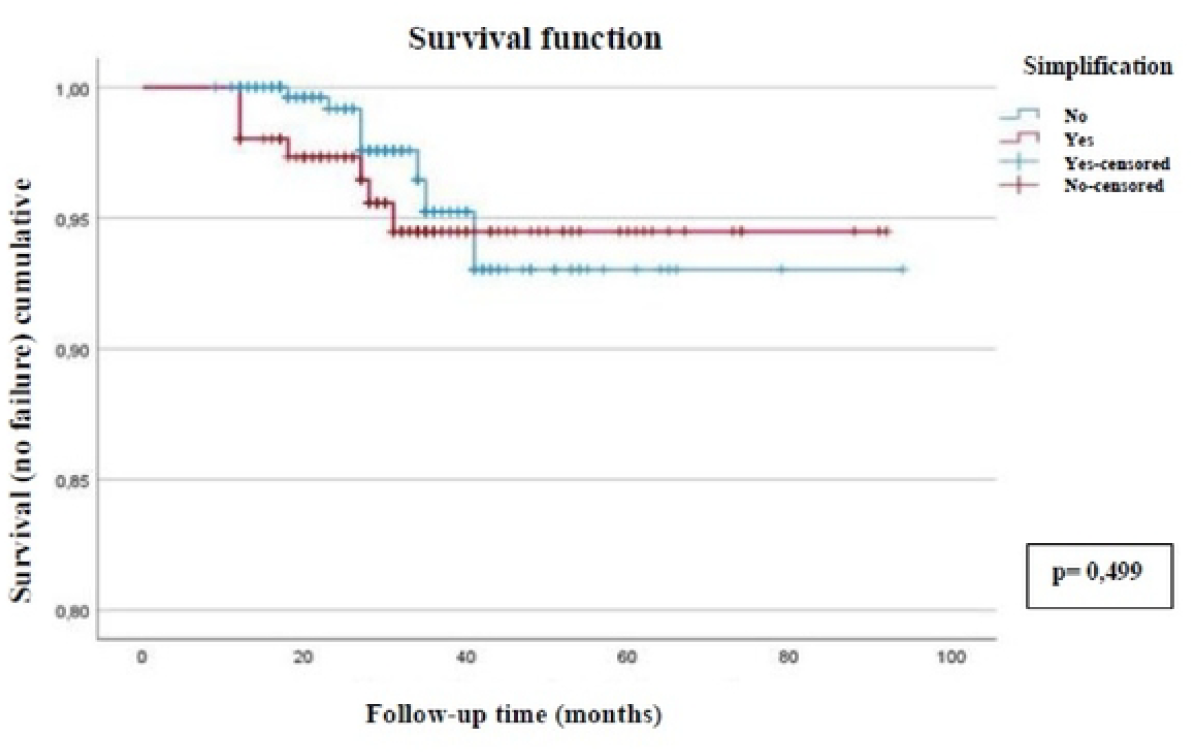
Kaplan-Meier curve illustrating therapeutic success (undetectable viral load) among 458 people living with HIV, comparing simplified and non-simplified antiretroviral regimens. Botucatu. Brazil. 2022.

At the end of follow-up, seven patients in the simplified group and eight patients in the non-simplified group had detectable VLs. This outcome was attributed to poor adherence or treatment dropout. The causes of poor adherence were primarily linked to psychological and social issues and not adverse events associated with ART.

The groups were heterogeneous in terms of the mean age (p < 0.001) and race (p = 0.0076). Regarding sex, the groups were homogeneous, as they had mostly male patients (p = 0.1972). Regarding the follow-up time, a difference was observed between the groups, as the simplified group had an average follow-up of 34 months, whereas the non-simplified treatment group was followed up for an average of 29 months (p < 0.001). The data are presented in Table 1.

**Table 1.** Demographic Characteristics of the 458 People Living with HIV (Botucatu, 2022)

| Variables | Simplified<br>(N = 152) | Non-simplified<br>(N = 306) | p-value |
| --- | --- | --- | --- |
| Age (years) | 54 | 42 | <0.001 |
| Sex, N (%) |  |  |  |
| Male | 96 (63.2) | 174 (56.9) | 0.1972 |
| Female | 56 (36.8) | 132 (43.1) |  |
| Race, N (%) |  |  |  |
| White | 134 (88.2) | 255 (83.3) | 0.0076 |
| Brown | 13 (8.6) | 16 (5.2) |  |
| Black | 5 (3.3) | 35 (11.4) |  |
| Follow-up time<br>(months) | 34 | 29 | <0.001 |

N: number
Student t-test

Among the 152 patients who underwent regimen simplification, 65 (42.7%) transitioned to a regimen with 3TC + DRV/r, while 87 (57.2%) transitioned to one with 3TC + DTG. The primary reasons for treatment simplification were osteopenia/osteoporosis, reduced eGFR, discontinuation of other NRTIs (ddI, AZT, or ABC), and other medical indications The category “other medical indications” included cases involving drug interactions, comorbidities, or concurrent use of medications with a potential risk of causing renal or bone toxicity, as detailed in Table 2.

**Table 2.** Characteristics of the 152 individuals living with HIV in Group 1 (patients with simplified antiretroviral regimens), categorized by the reasons for simplification (Botucatu, 2022)

| **Reasons for simplification** | **Simplified patients**<br>**(N = 152)**<br>**N (%)** |
| --- | --- |
| **Osteopenia/Osteoporosis** | 71 (46.7) |
| **Decreased eGFR** | 50 (32.9) |
| **Discontinuation of other NRTIs (ddI, AZT or ABC)** | 24 (15.8) |
| **Other medical indications** | 7 (4.6) |

Regarding the ART regimens before simplification, 46 patients (30.0%) were using 3TC + TDF + NNRTI, 42 patients (27.4%) were on 3TC + TDF + INI, and 40 patients (26.1%) were receiving 3TC + TDF + IP/r. Additionally, 25 patients (16.3%) were on regimens without TDF, consisting of 3TC combined with ABC, AZT, or ddI, along with IP/r or an NNRTI.

Among the 306 patients who maintained TDF, 169 (55.2%) received 3TC + TDF + INI, 86 (28.1%) received 3TC + TDF + IP/r, and 48 (15.6%) received 3TC + TDF + NNRTI. Only three patients (0.9%) were on regimens without TDF: ddI + 3TC + LPV/r, AZT + 3TC + DRV/r, and ABC + 3TC + DTG. Among these patients, a significant decrease in renal function (p < 0.001) was observed over a period of 29.4 ± 11.2 months.

In the simplified ART group (n = 152), 50 patients had their antiretroviral regimen simplified because of reduced renal function. An analysis of the eGFR variation in these patients revealed an improvement in renal function, demonstrated by a positive eGFR variation (final eGFR greater than the initial eGFR) over a follow-up period of 34.5 ± 14.8 months (Figure 2).

**Figure 2.**
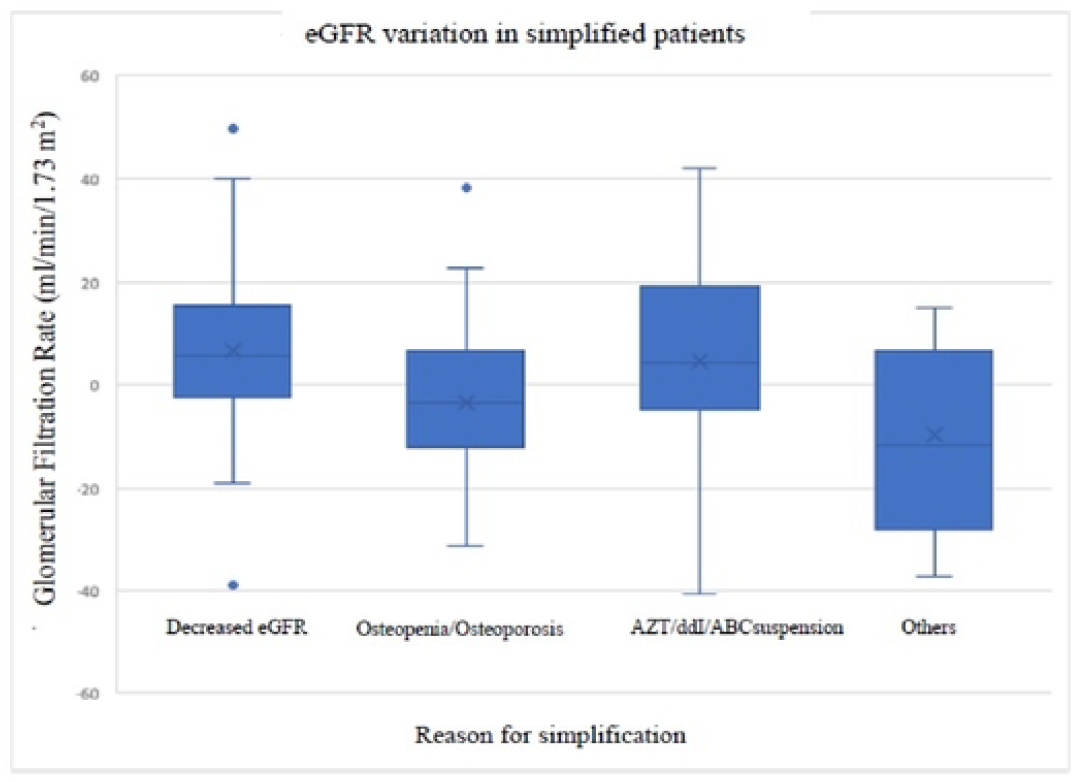
Variation in the estimated glomerular filtration rate among 152 people lwing with HIV on simplified antiretroviral regimens. categorized by the reason for regimen simplification. Botucatu, Brazil. 2022.

In the group with 152 simplified patients, 43 had osteoporosis or osteopenia as the reason for simplification. Of these 43 patients, 10 (23.3%) had an improvement in BMD, while 33 (76.7%) showed no change on examination during the follow-up period of two and a half years, on average, after simplification.

## 4. Discussion

This retrospective cohort study verified that simplified ART had the same effectiveness as triple therapy in relation to the maintenance of undetectable HIV VL, with no difference between the groups. This result is similar to that of other real-life studies that used 3TC+ DTG [15–19] or 3TC + DRV/r [15,20] as a strategy for simplifying ART and reducing side effects related to NRTIs, especially TDF.

Regarding patients who had a VL above the detection limit at the end of the follow-up period, the reasons were non-adherence and abandonment of therapy, as observed in most real-life studies, with no presence of resistance mutations [15–20]. The reasons for non-adherence and abandonment of therapy in the present study included psychological and social issues, and were not related to adverse events, according to data from medical records.

Unlike RCTs, the groups were heterogeneous in terms of several characteristics such as race, age, and follow-up time. Regarding the average age, patients who underwent simplification were older than those who did not. This probably occurred because losses in renal function and bone mass usually worsen with advancing age and longer exposure to TDF [21–23], and these were the main reasons for ART simplification.

One of the desired outcomes was an improvement in renal function after simplification. Therefore, the variation in eGFR was analyzed only in patients who had their therapy simplified because of renal dysfunction, and the result was a positive variation: an improvement in the final eGFR in relation to the initial one, within 34 months after the simplification. Some RCTs have shown improvements in renal function 48 weeks after discontinuing TDF [24,25]. In these studies, renal function was evaluated using eGFR calculations based on cystatin C levels in addition to the analysis of proximal tubule injury markers and the urinary protein/creatinine ratio [24,25].

In a prospective cohort study, Maggiolo et al. [26] observed a significant increase in serum creatinine levels two months after simplification in 94 patients simplified to a 3TC + DTG regimen. However, this increase was limited and stabilized after six months. This change aligns with the known mechanism of DTG as an inhibitor of the renal organic cation transporter 2, which mediates tubular creatinine secretion [26].

To better assess renal function following the discontinuation of TDF from the regimen, alternative parameters, such as serum cystatin C levels, proteinuria, urinary electrolytes, and urinary protein-to-creatinine ratio, may provide more accurate insights. Estimating creatinine clearance can be unreliable, as an increase in serum creatinine may reflect DTG-induced alterations in tubular secretion or increased muscle mass, rather than actual renal impairment, as suggested by Maggiolo et al. [26]. Cystatin C levels are not routinely assessed because of the high cost in Brazil; however, other urinary parameters can be assessed.

Although some studies have suggested eGFR improvement after discontinuing TDF, factors such as older age, longer duration of HIV infection, and comorbidities contributing to renal function decline have been associated with a reduced likelihood of complete renal recovery [21,25]. In the present study, comorbidities among PLHIV, such as diabetes, hypertension, and dyslipidemia, were not assessed, nor was the duration of HIV infection before regimen simplification. These aspects should be explored in future studies to provide a more comprehensive analysis.

Another outcome analyzed was BMD after simplification of the scheme, as some authors described an improvement in this alteration 48 weeks after discontinuing TDF [24,27]. The present study confirmed BMD improvement in 23.3% of patients with osteoporosis or osteopenia, and BMD maintenance in most of them during the analyzed period, an average of two and a half years after simplification. Of note, the BMD of patients did not worsen, which could probably have happened if they maintained the use of TDF.

An alternative method of evaluation involves the analysis of bone remodeling biomarkers such as bone alkaline phosphatase, serum osteocalcin, C-terminal telopeptide, and type 1 collagen propeptide. Evidence from two RCTs demonstrated that these markers reduced in regimens excluding TDF, indicating decreased bone loss [24,27]. However, these biomarkers were not assessed in our study.

In the RCTs reviewed, comparisons between groups revealed improvements in BMD patterns and bone resorption markers among patients who transitioned to simplified regimens, compared with those who continued TDF-based regimens [24,27]. In contrast, the present study evaluated only the simplified group, precluding direct comparisons between groups.

Despite some limitations, such as the demographic heterogeneity of the groups and the relatively small sample size, the results of the present study are robust and consistent with those of other real-life studies and RCTs. Furthermore, this was a pioneering study comparing two groups of patients (simplified and non-simplified) in an observational manner.

More real-life studies are needed to prove the effectiveness of simplified therapy in maintaining viral suppression and verify the benefits in relation to bone and renal health. In addition, further studies are needed to determine the best moment to perform ART simplification to avoid these changes.

## Data Availability

http://hdl.handle.net/11449/243161

## Ethical Approval Statement

This study was approved by the Research Ethics Committee of Botucatu Medical School– UNESP (State University of Sao Paulo) on July 5th, 2022, under the ethical approval reference number:59638222.0.0000.5411.

